# Monitoring the opioid epidemic via social media discussions

**DOI:** 10.1101/2021.04.01.21254815

**Authors:** Delaney A. Smith, Adam Lavertu, Aadesh Salecha, Tymor Hamamsy, Keith Humphreys, Mathew V. Kiang, Russ B. Altman, Johannes C. Eichstaedt

## Abstract

Opioid-involved overdose deaths have risen significantly since 1999 with over 80,000 deaths annually since 2021, primarily driven by synthetic opioids, like fentanyl. Responding to the rapidly changing opioid crisis requires reliable and timely information. One possible source of such data is the social media platforms with billions of user-generated posts, a fraction of which are about drug use. We therefore assessed the utility of Reddit data for surveillance of the opioid epidemic, covering prescription, heroin, and synthetic drugs (as of September 2024, up-to-date Reddit data was still accessible on the open web). Specifically, we built a natural language processing pipeline to identify opioid-related comments and created a cohort of 1,689,039 geo-located Reddit users, each assigned to a state. We followed these users from 2010 through 2022, measured their opioid-related posting activity over time, and compared this posting activity against CDC overdose and National Forensic Laboratory Information System (NFLIS) drug report rates. To simulate the real-world prediction of synthetic drug overdose rates, we added near real-time Reddit data to a model relying on CDC mortality data with a typical 6-month reporting lag and found that Reddit data significantly improved prediction accuracy. We observed drastic, largely unpredictable changes in both Reddit and overdose patterns during the COVID-19 pandemic. Reddit discussions covered a wide variety of drug types that are currently missed by official reporting. This work suggests that social media can help identify and monitor known and emerging drug epidemics and that this data is a public health “common good” to which researchers should continue to have access.

**Significance statement:** The opioid epidemic persists in the United States with over 80,000 deaths annually since 2021, primarily driven by synthetic opioids like fentanyl. As the geographic and demographic patterns of the opioid epidemic are rapidly changing, accurate and timely monitoring is needed. In this paper, we used social media data from Reddit to conduct public health surveillance of the opioid epidemic, following 1.5+ million geo-located users over 10+ years. We also found that near real-time Reddit data can improve our ability to predict future overdose death rates compared to models only using CDC data with typical half-year reporting delays. Our work suggests that social media can be a useful component for public health surveillance of the opioid epidemic.

## Introduction

The United States is currently experiencing an opioid epidemic. The number of opioid-related deaths has been rising almost every year since 1999, increasing significantly in 2021 and 2022.^1^ Much of this increase has been driven by synthetic opioids, such as illicitly manufactured fentanyl and its analogs. Fentanyl is 50 times stronger than heroin and 100 times stronger than morphine.^1^ Overdose rates from synthetic opioids have increased by over 22% from 2020 to 2022, with the 2021 annual rate of 73,838 deaths per US population being 22 times greater than the 2013 rate.^1^ To facilitate a more effective response to this epidemic, public health agencies must be able to quickly identify, monitor, and address both established and emerging patterns of non-medical use.^2^

To that end, the U.S. spends billions of dollars every year to conduct public health surveillance, and this surveillance relies on the integration of data from many different sources. For instance, the National Institute on Drug Abuse (NIDA) has several existing systems for tracking rates of non-medical drug use at the population level. NIDA’s National Drug Early Warning System (NDEWS) integrates information on drug use from community epidemiologists at satellite offices around the country.^3^ NIDA’s Monitoring the Future effort conducts annual surveys of thousands of students from hundreds of schools, asking questions about drug use experiences in the recent and distant past. The National Vital Statistics program at the CDC collects data on deaths from a large number of regional entities within the United States, identifying causes of mortality, including drug overdoses.^4^ The National Forensic Laboratory Information System (NFLIS) is a DEA program that monitors national drug patterns through reports from state and local forensic labs.^5^ The Substance Abuse and Mental Health Services Administration (SAMHSA) in the U.S. Department of Health and Human Services (HHS) leads public health efforts to reduce harm related to substance use disorders.^45^

These systems are accepted by research and policy communities; they continue to grow in scope and have established benchmarks, methods, and resources for analysis. Despite these benefits, these systems have limitations in monitoring ongoing and developing drug epidemics. Specifically: (1) they are not real-time, (2) some survey outcomes are subject to interpretation and rely on an expert’s perception of another person’s health (i.e., epidemiologists, doctors, social workers), (3) they are sparse in terms of geographic, demographic, and/or disease coverage, (4) are constrained in their ability to discover novel and unexpected trends, and (5) they rely on data collection methods that can be biased.^6^ For example, a recent study found that the National Survey on Drug Use and Health (NSDUH) general population survey of 70,000 participants was inadequate for monitoring heroin use, and describes many of the above limitations.^8^

The U.S. Food and Drug Administration (FDA) and others have identified social media as a potentially useful source of real-world evidence for drug monitoring.^7,9^ However, public health efforts have been slow to adopt social media surveillance due to challenges currently posed by the data. Social media datasets can be very large and most of their content is not relevant to public health, thus, it requires additional expertise and resources to mine and analyze the data.^10^ Additionally, evidence of the utility of social media data is still relatively limited compared to evidence for traditional methods of public health surveillance.

Social media could offer unique benefits to epidemic monitoring, such as access to an observational data source that is potentially uncorrelated with the data streams currently used in public health surveillance. Moreover, the pseudo-anonymous nature of these platforms can make some users more inclined to discuss stigmatized behaviors and their health and health issues.^1,9^ The data are mostly first-person reports, thus not potentially biased by expert interpretation. Users’ posts often include additional details about their lives or environments providing contextual information often lacking from other reporting systems. Finally, social media data is both retroactively and rapidly available, potentially providing a robust longitudinal data set to identify trends.

The majority of research on drug and disease surveillance in social media has focused on Twitter (recently renamed X), although some research has used Reddit, Facebook, Instagram, or smaller online discussion forums.^12^ A review article by Sarker *et al*. found over 1,000 articles since 2012 related to social media and drug use monitoring.^13^ In 2018, Harrigian attempted the first geolocation of Reddit posts using text-based heuristic schema with 45% accuracy on US data.^14^ Narrowing the focus to geolocated drug monitoring, Katsuki *et al*. found an association between prescription non-medical drug use and illicit online pharmacies on Twitter.^15^ Chary *et al*. used a set of keywords to scan Twitter for opioid mentions and filtered the resulting mentions based on semantic distance to a set of manually labeled tweets. These tweets were grouped by geolocation, and the total count for each state was normalized by the total number of tweets collected from that state. They found a strong correlation between normalized Twitter frequency values and opioid use statistics from the NSDUH.^16^ A recent study by Sarker et al. followed 9,006 Twitter posts from Pennsylvania and demonstrated that machine learning could derive drug use tweet rates that were correlated with county-level opioid overdose death rates.^17^ Studies that specifically use Reddit data include a study by Pandrekar *et al*. that investigated the opioid discussion on Reddit by analyzing 51,537 opioid-related posts, performing topic modeling, and finding the psychological categories of the opioid posts.^18^ Chancellor *et al*. used Reddit posts about opioid recovery and discovered potential alternative treatments in opioid recovery posts.^19^ More recently, Garg *et al*. determined individual risk levels of fentanyl use using a machine-learning model trained on Reddit data.^20^

There is an opportunity to use data from the social media platform Reddit to conduct public health surveillance of the opioid epidemic, and to benchmark its value as an input to a time series prediction system. Reddit, a pseudo-anonymous platform, was founded in 2005 and has been in the top ten most-visited websites in the United States for over ten years.^20^ User activity on the Reddit platform occurs in topic-oriented subreddits, denoted with a leading ‘r/’, where users can congregate to discuss a particular topic of interest. Community moderators and users work to keep discussions focused on particular topics. For example, ‘r/addiction’ is a subreddit where community members discuss and support each other in overcoming their addictions. In 2022, there were over 57 million daily active users on Reddit, who contributed over 422 million posts, and 2.86 billion comments.^22^ Unlike other social media platforms like Twitter and Facebook, the majority of the historical activity on Reddit was archived and is publicly available.^23^ Overall, 11% of adults in the U.S. report using Reddit.^24^ The user base is skewed towards males, younger individuals, White individuals, those of Hispanic descent, and those that have at least some college education^24^. Although these demographics differ from those of the United States overall and would be limiting in the study of drugs used primarily in older populations, they may be sufficient for monitoring the rate of drug use in represented groups.

In this study, we create a cohort of 1,689,039 users and follow them over a period of 10+ years, measuring drug-related discussion over time. We create an approximate geo-location algorithm to map users to their likely locations. We examine the overall discussion rate of opioids on the Reddit platform, including that of individual drugs and drugs grouped by opioid class. We demonstrate that our social media-derived synthetic opioid trends are comparable to both CDC overdose death rates and NFLIS drug report rates across national, and regional analyses. We also examine the utility of Reddit data in predictive modeling of overdose death rates. We focus on the relatively recent rise in discussions of synthetic opioids, including fentanyl, a powerful and deadly synthetic opioid that has risen in use in the United States. Finally, we examine recent changes during the COVID-19 pandemic. Our work demonstrates the utility of monitoring social media for conducting timely surveillance of the opioid epidemic.

## Results

### Summary statistics

Our final comment dataset included 6,344,026 opioid mentions from 6,065,600 comments. Heroin (and synonyms from the RedMed drug term lexicon) is the most frequently mentioned opioid on Reddit, followed by morphine, fentanyl, and oxycodone (Table 1).^35^ We categorized mentions by opioid class. Opioids are either naturally occurring or synthesized and act as either agonists or antagonists. Opioids that are agonists have the highest prevalence of non-medical use.^5^ Fentanyl was the most frequently mentioned agonist and Naloxone was the most frequently mentioned antagonist. Partial agonists and mixed agonists were mentioned less.

**Table 1.** Opioid mentions, unique opioid comments, and unique users mentioning opioids on Reddit between January 2006 and December 2022. Heroin has more than four times the number of comments of the next most talked about opioid (Morphine) and is mentioned by the broadest number of users. The number of mentions quickly decreases for lesser-known opioids, especially those of the mixed agonist class (e.g., Nalbuphine).

| Drug | Opioid Class | Mentions | Comments | Users |
| --- | --- | --- | --- | --- |
| Heroin | Heroin | 2,087,560 | 2,048,960 | 1,473,202 |
| Morphine | Full Agonist | 457,885 | 435,300 | 328,491 |
| Fentanyl | Full Agonist | 393,993 | 370,313 | 262,710 |
| Oxycodone | Full Agonist | 359,244 | 363,869 | 260,609 |
| Naloxone | Antagonist | 362,160 | 345,618 | 203,306 |
| Hydrocodone | Full Agonist | 306,704 | 302,852 | 224,750 |
| Methadone | Full Agonist | 282,839 | 268,525 | 154,799 |
| Codeine | Full Agonist | 216,807 | 210,109 | 152,371 |
| Tramadol | Full Agonist | 157,086 | 144,402 | 96,556 |
| Hydromorphone | Full Agonist | 101,425 | 99,344 | 70,813 |
| Buprenorphine | Partial Agonist | 97,798 | 92,856 | 58,277 |
| Loperamide | Partial Agonist | 84,013 | 82,936 | 61,082 |
| Naltrexone | Antagonist | 72,317 | 70,679 | 43,845 |
| Pethidine | Full Agonist | 13,884 | 13,328 | 10,599 |
| Nalmefene | Antagonist | 1,074 | 5,278 | 3,659 |
| Nalbuphine | Mixed Agonist | 1,942 | 1,834 | 1,383 |
| Sufentanil | Full Agonist | 1,812 | 1,771 | 1,395 |
| Butorphanol | Mixed Agonist | 1,585 | 1,535 | 1,201 |
| Pentazocine | Mixed Agonist | 1,075 | 1,018 | 823 |
| Levorphanol | Full Agonist | 478 | 439 | 340 |
| Diprenorphine | Antagonist | 526 | 337 | 262 |
| Nalorphine | Mixed Agonist | 55 | 51 | 43 |
| Dezocine | Mixed Agonist | 47 | 36 | 26 |

### Cohort statistics

We mapped 1,689,039 unique users to 46 states by exploiting the fact that some Reddit users have also posted in location-specific subreddits (e.g., r/Philadelphia). Within this cohort, 258,591 users mentioned an opioid on Reddit at some point in their comment history. Kendall’s Tau correlation between a state’s estimated 2020 population and our number of observed users was 0.595 (see Supp. Figure 1). We followed this group from 2009 to December 2022, during which time they posted about 1.7 billion comments on Reddit.

### Comparisons with benchmark opioid statistics

#### Benchmark: CDC vital statistics overdose data

We compared 12-month trailing (that is, aggregation across a 12-month rolling window) U.S. opioid comment rates on Reddit against 12-month trailing opioid overdose death rates for different geographies (U.S., region) and drug categories (see Figure 2, left). We explored lead/lag differences between the two time series using cross-correlations for different lag offsets (Figure 2, right). We found similar trends between Reddit comment rates and overdose death rates, especially for synthetic opioids (with a simple time series cross-correlation of r = 0.89 with no lag) (Figure 2, top). To guard against spuriously elevated correlations of time series data, we detrended time series (first-order differencing), resulting in a cross-correlation of r = 0.59, which was still the highest with no lag. Consistency of distributions was confirmed through the augmented Dickey-Fuller unit root test, p < 0.001) for synthetic opioids in both CDC and Reddit time series, meaning that correlations between the time series remained after detrending.

**Figure 1.**
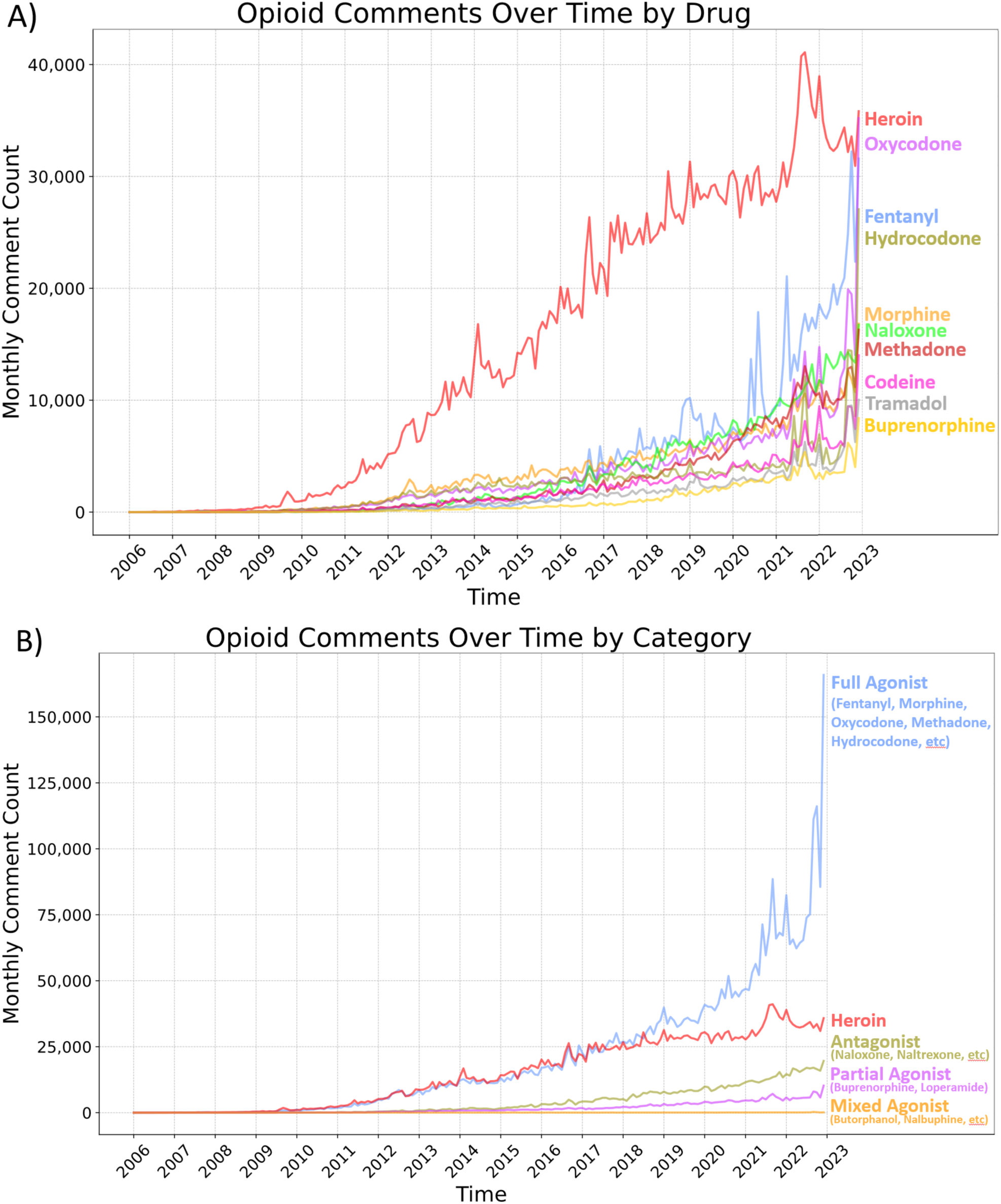
Growth of opioid discussions amongst all users on Reddit. **A)** The discussion about opioids on Reddit over time for the top 10 most discussed opioids. Fentanyl has become the most frequently mentioned non-heroin opioid in recent months. **B)** Comment counts for opioids categorized by mu-opioid receptor activity. Mentions of full agonist opioids, those with the highest pain relief and non-medical use potential, have been on the rise since 2010 at a rate similar to heroin mentions and fully surpassing heroin mentions by mid-2017. This change in rates is not identifiable from analyzing the individual opioids.

**Figure 2.**
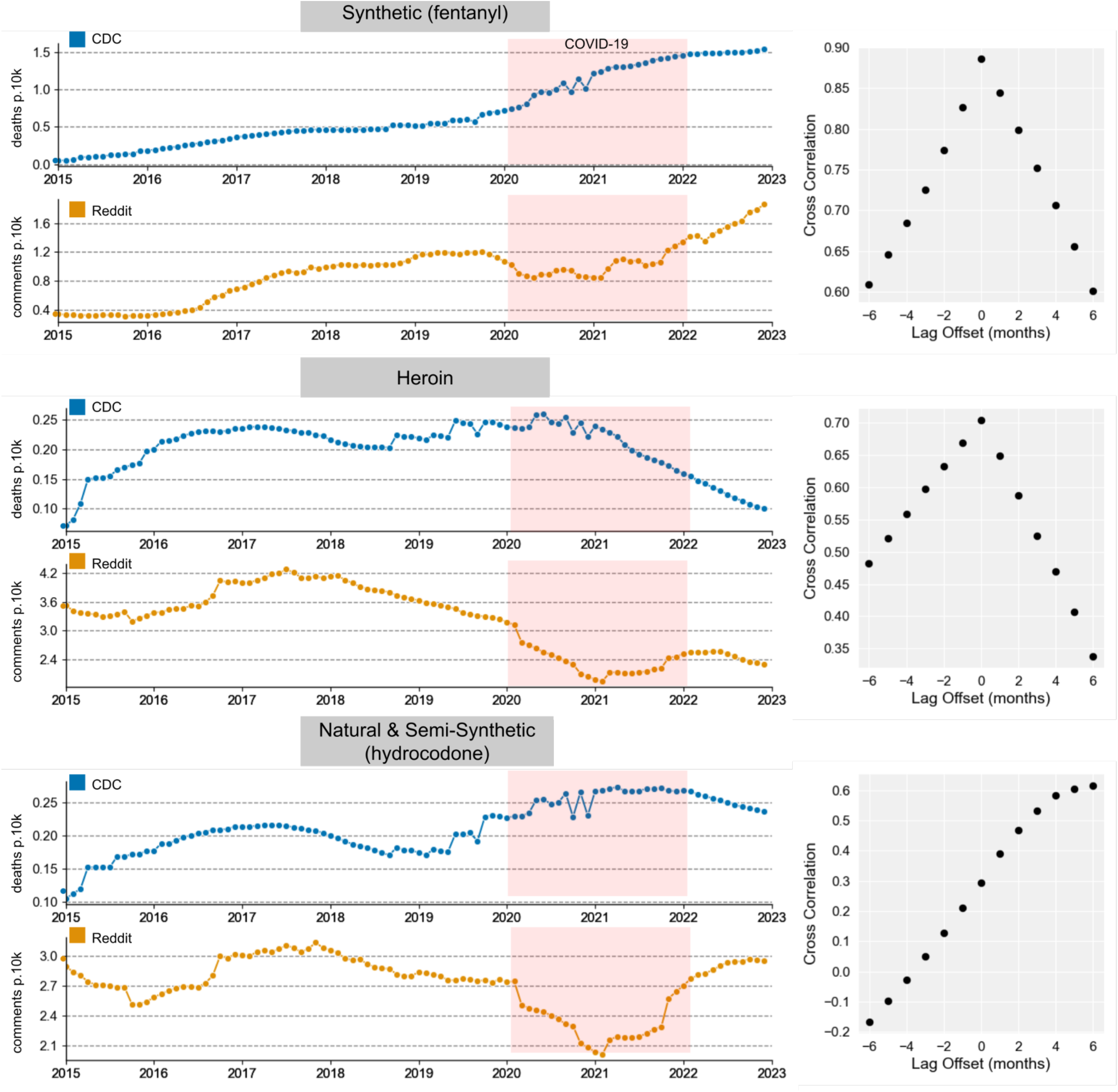
**Left**: 12-month trailing monthly U.S. opioid overdose CDC death rates per 10,000 people (top panel of subplots, in blue), and monthly opioid comment rates on Reddit per 10,000 total comments (bottom panel of subplots, in orange) for different drug categories, along with **Right**: the results of 6-month lead/lag cross-correlation analysis. The cross-correlation analysis indicates that for synthetic opioids, a lead/lag of zero yields the best correlation, indicating that while the magnitude of changes may differ, the shape of the Reddit comment rate over time vs. the benchmark over time is relatively synchronous. The same trend is observed for heroin. Natural and Semi-synthetic CDC trends lag behind Reddit trends by about 5 months. The COVID-19 pandemic is shaded in red from January 2020 to December 2021 and is excluded from the cross-correlation analysis.

We also examined heroin and natural/semi-synthetic opioids and found similar trends for all years except 2020 and 2021. In general, Reddit posts seemed to be less accurate proxies for these substance classes than they were for synthetic opioids. Heroin had a correlation value of r = 0.71 with no lag, while Natural and Semi-synthetic opioids had a correlation of r = 0.62 when CDC rates trailed Reddit rates by five months (Figure 2). After de-trending (first-order differencing), the correlation values became r = 0.60 for Heroin and r = 0.25 for natural and semi-synthetic drugs (stationarity was confirmed through augmented Dickley-Fuller tests in all cases, p’s < 0.05). While r values lowered after de-trending, the remaining time series were still positively correlated.

#### Benchmark: NFLIS data

We compared Reddit opioid activity with National Forensic Laboratory Information System (NFLIS) drug report rates over time, which are semi-annual published reports that provide the estimated number of total drug reports submitted to laboratories with region-level geographic resolution (Figure 3). In particular, we compared semi-annual Reddit opioid activity with semi-annual NFLIS drug report rates for 2014-2022 for fentanyl, heroin, and hydrocodone as other key drugs for the opioid epidemic. We compared rates over time at the national level and across the Midwest, West, South, and Northeast regions. For fentanyl, in particular, we observed Reddit trends to match NFLIS trends across all regions. All trend lines show divergence between Reddit and NFLIS during the COVID-19 pandemic in 2020 and 2021, which appeared to resolve by 2022.

**Figure 3:**
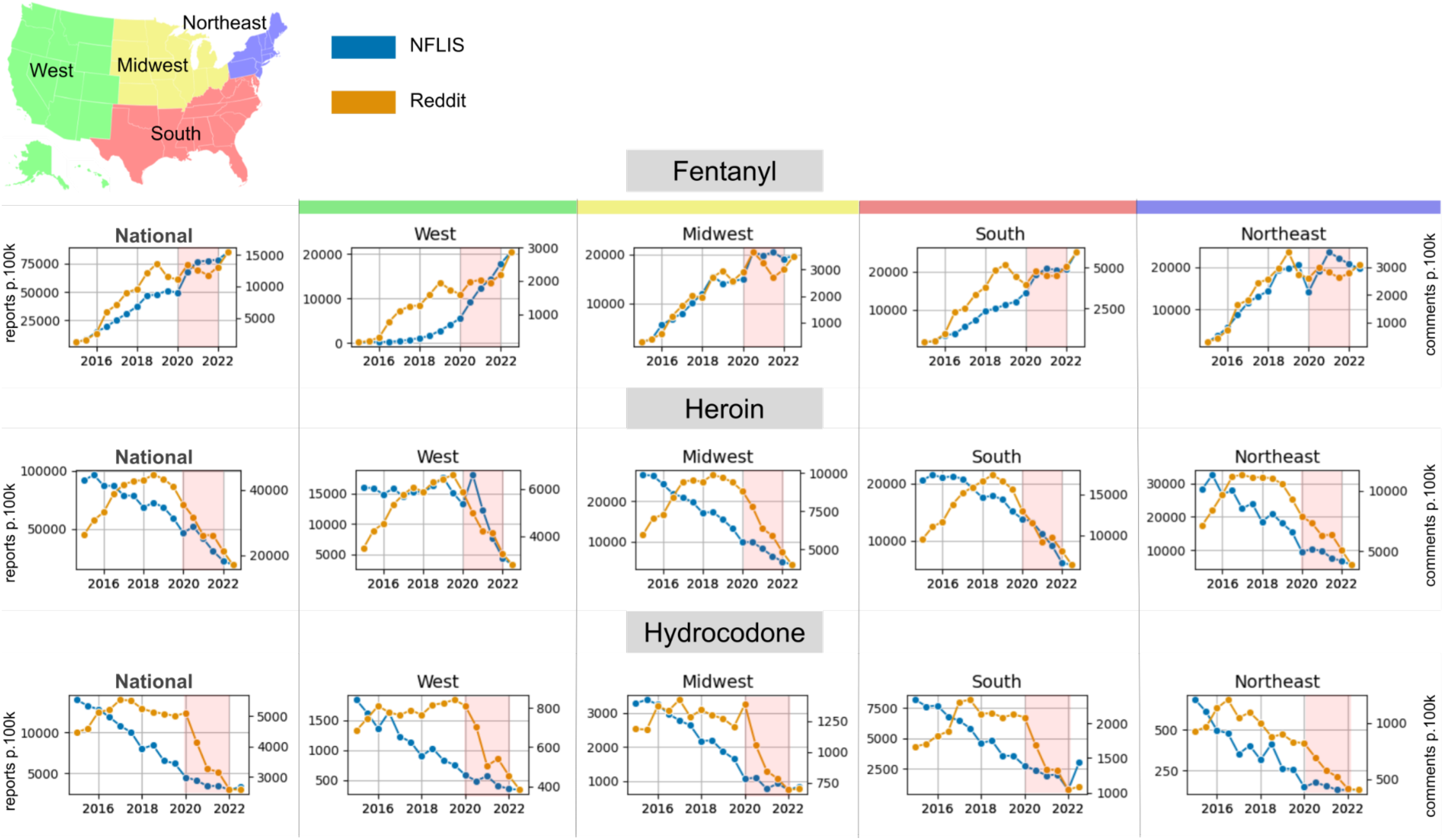
Fentanyl (top), heroin (middle), and hydrocodone (bottom) semi-annual NFLIS opioid report rates per 100,000 people (orange line, left axes) and semi-annual Reddit opioid comment rates per 100,000 total comments (blue line, right axes) from 2014 to 2022 for, plotted at the national (left) and regional levels. We observe similar trends between Reddit and CDC rates for all drugs except during the COVID-19 pandemic, shaded in red from January 2020 to December 2021.

Reddit and NFLIS time series for fentanyl correlated at r = 0.91; after detrending through first-order differencing, this reduced to r=0.41, suggesting similarity of the time series even after the general trends were removed (stationarity confirmed through an augmented Dickey-Fuller test, p < 0.05). For heroin and hydrocodone, Reddit and NFLIS time series correlated at r = 0.63 and r = 0.77, respectively, but could not be detrended (stationarity could not be achieved using first or second-order differencing transformations).

### Time Series Predictive Modeling

In real-world applications, CDC mortality estimates are available with at least six months of delay and thus are only available for prediction of future mortality with such a lag. Social media data, however, is available in near real-time, which may yield increased predictive accuracy when added to the lagged CDC data. Thus, we assessed the utility of the Reddit data at the national level in predicting CDC overdose death rates using an autoregressive integrated moving average model (ARIMA) from 2015 through 2022. We used rolling origin forecasts to estimate the absolute error at 1-month prediction horizons of overdose death rates. We compared the performance of 6-month lagged CDC data alone and in combination with 1-month lagged Reddit data in forecasting monthly overdose death rates per 10,000 people (see Figure 4A for method summary). We found that adding near real-time Reddit data as an exogenous variable significantly improved the model’s prediction accuracy at the national level for synthetic opioids (Wilcox signed rank test of absolute errors, p = 0.019) (Figure 4B). We also report the comparative distribution of absolute errors for both models (Figure 4C, D). The combination Reddit/CDC model had more instances of lower absolute errors than the CDC model alone. The average absolute error for CDC alone was 0.0287, and 0.0246 in normalized monthly opioid overdose deaths (per 10,000 people) for the combination model. In both models, we see notably decreased performance during the COVID years from 2020 until 2022.

**Figure 4.**
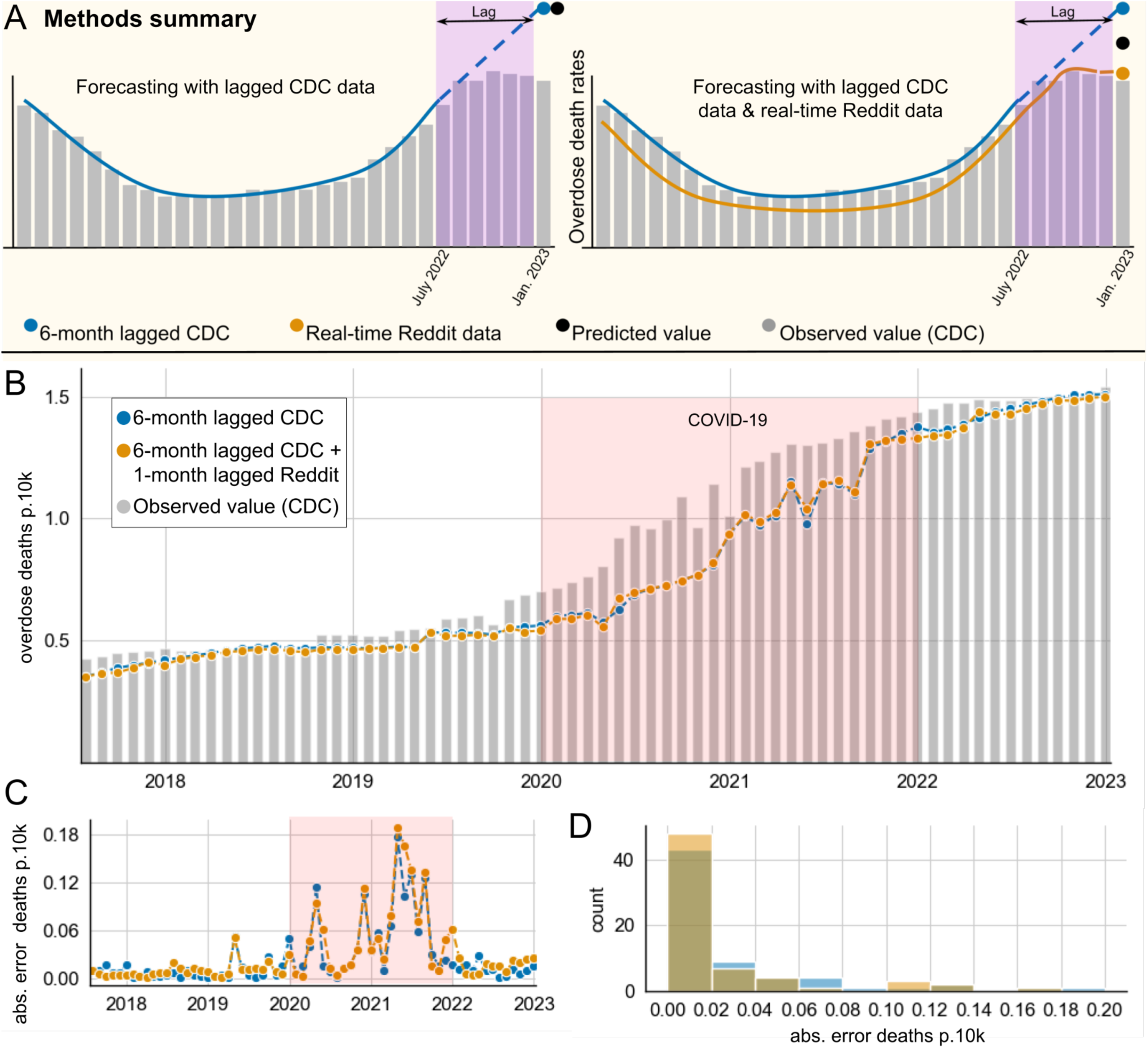
Autoregressive Integrated Moving Average (ARIMA) simulations for synthetic opioids**. A)** Methods summary. As an example, to obtain the mortality rate estimate for January 2023, we combined 6-month lagged CDC data (blue trendline) with 1-month lagged Reddit data (orange trendline). Grey bars show the observed monthly overdose death rates per 10,000 people as reported by the CDC once available; solid lines represent data incorporated into the prediction, but dashed line data is not included. In this example, the predicted overdose death rate for January 2023 is generated by CDC data from July 2022 alone (left), or in combination with Reddit data from December 2022 (right). **B)** Predicted monthly overdose death rates per 10,000 people of rolling-origin forecast models are shown based on 1-month prediction horizons. Observed mortality is shown in grey, and monthly overdose death rates per 10,000 people as predicted by ARIMA models fitted on 6-month-lagged CDC overdose death (blue) are shown along with predictions from a model that additionally included 1-month-lagged Reddit data (orange). **C)** shows the absolute errors over time of the monthly overdose deaths per 10,000 people predicted by the lagged CDC model alone (in blue) and the combined CDC/Reddit model (in orange); **D)** shows the corresponding distributions.

## Discussion

Since 2013, synthetic opioids like fentanyl have emerged as a major contributor to the opioid epidemic and the total number of opioid deaths.^1^ The COVID-19 pandemic made the opioid epidemic worse, especially among those with pre-existing social, economic, and health disadvantages and potentially due to unemployment and isolation as a result of COVID-19 policies.^25–27^ The ubiquity of social media platforms can provide a lens into the drug use of millions of Americans. The continued growth of opioid-related discussions on Reddit demonstrates that social discourse around drug use is expanding online (Figure 1). In this paper, we benchmark a method for monitoring the online social footprint of drug epidemics spatiotemporally using Reddit. We find that Reddit can be a useful, alternative data stream as the number of opioid mentions (across all linguistic contexts) correlates robustly with other opioid statistics while also providing spatial granularity, long-term continuity, and additional ethnographic information that contextualizes drug mentions.

We found that Reddit comment rates have risen in sync with overdose (CDC) and laboratory (NFLIS) rates at both the national and regional levels and show strong correlations with CDC overdoes even after de-trending. The combined near real-time Reddit synthetic opioid ARIMA model had significantly better accuracy in predicting overdose death rates at the national level compared to the lagged CDC-only model, suggesting Reddit data provides incremental predictive validity.

Specifically, we observed that the Reddit comment rates for synthetic opioids in particular closely follow the CDC benchmark rates over time at the national level (simple correlation with CDC data: r = 0.89: detrended: r = 0.59) (Figure 2), with similar trends for heroin and less signal for natural and semi-synthetic opioids. For synthetic opioids and heroin, Reddit trends tend to match CDC trends best with no lag. We generally observed strong correlations between CDC and Reddit trends over time (r = 0.60 to r = 0.89), which reduced to r = 0.25 – 0.60 after trend differencing, suggesting that some of the measured correlations were due to linear trends in the data. However, the fact that stationary cross-correlations remained significant demonstrates that Reddit captures drug trend data over and above broad overall trends, and points to the usefulness of Reddit as an alternative data stream to monitor the opioid epidemic.

Beyond broad drug categories reported by the CDC, we used data from the National Forensic Laboratory Information System (NFLIS), which reports only semi-annually but reports individual drugs. We found Reddit mentions of fentanyl to be highly similar to semi-annual NFLIS fentanyl laboratory data at both the regional and national levels (r = 0.91, detrended: r = 0.41; Figure 3). We also observed heroin and hydrocodone comment rates to roughly match NFLIS benchmarks. This robust pattern of correlations indicates that Reddit surveillance may capture the use of single drugs, especially at the national level. This range of drug coverage on Reddit may allow for the monitoring of other drugs of interest that are not currently monitored by the NFLIS, including emerging drugs.

We wanted to assess how incorporating Reddit data might improve real-world drug overuse forecasting. We hypothesized that social media may provide more frequent and more recent trend information than traditional monitoring systems such as the CDC Vital Statistics Provisional Drug Overdose data, which often lags by at least four months.^42^ However, the wait time is much longer (sometimes years) for finalized overdose drug data to be released by the CDC. To model this scenario where delayed CDC estimates were available, but Reddit data was up to date, we created an ARIMA model for predicting overdose death rates in which the CDC data lagged by six months. We then compared the performance of this model to an ARIMA that included near real-time (1-month lag) Reddit mention rates as an exogenous variable (Figure 4) and observed significant improvement in prediction accuracy of overdose death rates from synthetic opioids. This demonstrates the utility of social media data to predict synthetic opioid trends in near real-time using a combination of social media and traditional reporting systems, and points to the potential of future systems to build on this work.

These prediction accuracies matter: Fentanyl overdoses have been rising steadily and timely response can save lives.^1^ Fentanyl is very potent, often undetectable, and can be laced into other recreational drugs. Fentanyl overdose can be treated using fast-acting opioid antagonists like naloxone, which need to be allocated and distributed as quickly as possible. Our results suggest that Reddit data may become part of an early-warning system that predicts geographic changes in fentanyl use, allowing harm reduction programs to adequately stock and distribute naloxone and deploy drug testing strips and other public health interventions.

In contrast, we observe that incorporating Reddit data into predictive models for heroin and natural/semi-synthetic opioids did not yield significant improvements in predictive accuracy. This may be because both heroin and natural/semi-synthetic opioid trends have been stable or decreasing over the period we were assessing. We also attempted to achieve single-drug resolution in ARIMA models using NFLIS data. However, since NFLIS data is only reported semi-annually, we found that this increased forecast horizon dramatically reduced the prediction performance of the ARIMA models. This demonstrates that the frequency of data collection impacts the quality of the time series models and points to the value of near real-time social media data as a data source.

Using social media as a proxy for opioid overdose rates proved challenging during the COVID-19 pandemic. While societal changes resulted in unemployment and social upheaval, we observed that overall Reddit opioid comment rates increased dramatically (Figure 1). In terms of a normalized rate metric in which opioid mentions are divided by the total number of comments (as seen in Figures 2 and 3), we observed the rate to plateau or drop during 2020 and 2021, as other non-opioid discourse ‘drowned out’ opioid mentions. In contrast, the CDC observed that the rate of opioid overdoses drastically accelerated during the COVID-19 pandemic (Figure 2, top). Reddit comment rates re-aligned with benchmark trends in 2022 after many pandemic measures had been lifted (and presumably, Reddit discourse had normalized). These patterns suggest that our method, like many other monitoring systems, is not robust to large-scale behavioral changes and major systemic shocks such as the pandemic.^28^ In our time series predictive modeling, we also saw that all models performed better pre-COVID (before 2020). This shows that while Reddit data improved models that included pandemic data, this system is more useful when trends are not affected by global societal upheaval.

Our approach employed a relatively simple location mapping strategy. It is based on the assumption that Reddit users are most likely to comment on the location subreddit for their own location, not accounting for people who move cities or post in multiple location subreddits –incorrect location information adds noise to our results. Although this assumption may be reasonable at aggregate levels, the creation of geo-located cohorts with more sophisticated location mapping techniques could yield an improved drug monitoring system. Combining Reddit data with other social media data streams, such as X (Twitter), could further improve our results, and techniques have evolved to geolocate X users down to the county level.^43^ Moreover, we do not perform a fine dissection of the linguistic content or demographic information surrounding opioid mentions. We elected to build our model using opioid mentions alone because both folks with opioid use disorder and other affected groups may discuss opioids in a variety of ways, and our rate metric sums across all these mentions. Demographic information is not directly available on Reddit. However, additional linguistic analysis and subreddit analysis can be used to infer location, interests, gender, and other demographic features of interest. This is of interest because the demographic of Reddit users differs from the US population, leaning towards younger male users.

Monitoring systems such as ours depend on the continuing availability of data, which has been changing in recent years. Before late 2023, Reddit data could be easily accessed through the PushShfit Application Programming Interface (API).^44^ This was advantageous for a near real-time warning system. However, the PushShift API ceased to be available, and the official Reddit API, although more limited, would be an alternative for accessing new data. However, as of September 2024, up-to-date Reddit archives are circulating on the open web and can be accessed by researchers. Twitter accessibility has also changed since 2022, with more stakeholders asking for access for public health and civic purposes. Our work stresses the importance of social media data as a “common good” for public health and adds force to the demands made by the US Surgeon General in May 2024 and the regulation passed by the EU parliament that requires researchers to be given access to the data of large social media platforms.^39, 40^

There are critical ethical considerations when using online discussions to monitor non-medical drug use, including ensuring that user anonymity is preserved. While Reddit is a pseudo-anonymous platform (some handles contain no identifying information, whereas others may be identifying to some degree), we do not share any usernames in this study, and we do not analyze any user on the individual level, in accordance with ethical guidelines.^29^ Our study relies on the creation of a cohort that we have followed for more than a decade, and all analyses are performed aggregated by location. A formal system implemented to monitor social media data continuously with higher spatial resolution would have to balance protecting user anonymity and societal benefit from improved drug use surveillance.

If social media surveillance came to greater public awareness, people who use drugs might change their posting behavior and thereby change the reliability of our results. For example, the community could find or create new private communication platforms. Indeed, there have already been conversations on Reddit about moving such discussions to private chat rooms (such as Telegram and WhatsApp), which would hinder surveillance by third parties.^30^ The degree to which these discussions occur in public versus private domains will be a key determinant of the performance of a potential future surveillance system.

A key limitation of this study is that we do not distinguish between drug mentions and probable drug use (or any behavioral context) when computing drug mention rates. Our results suggest that drug mention frequency is a reasonable indicator of drug use in a region, regardless of why the drugs are mentioned – we have not established that these drug mentions are coming from drug users or their close contacts. It is also conceivable that news cycles impact rates of Reddit drug mentions, but we did not observe such patterns in the data.

Further, the way drugs are referenced on social media is continuously evolving, which the research community is tracking. For example, a recent work by Alhamadan *et al*. uses the r/drugs and r/opiates subreddits to recognize evolving drug terms.^31^ In addition, the increasing sophistication of Large Language Models (LLMs) may be helpful in assessing the likely context of drug mentions at scale, which may improve accuracy and provide contextual information, which we are exploring in current research. An advantage of the high volume of drug discussions on Reddit combined with the availability of LLMs to streamline Natural Language Processing tasks means that systems can be built relatively cost-effectively and quickly to spot emerging drug trends that are not yet monitored by the CDC or NFLIS – a true early warning system.

In conclusion, we introduce a cohort-based approach for monitoring geographical rates of opioid mentions on Reddit over time, which corresponds reasonably well to established official mortality and laboratory monitoring systems. We develop time-series forecasting models using both CDC and Reddit data, showing that Reddit data adds predictive power to these models, especially for synthetic opioids, such as fentanyl. Social media has the potential to provide near real-time information about evolving drug use trends across a wide variety of known and emerging drugs, and it contains additional ethnographic information that can be used to contextualize drug use. More generally, our work demonstrates that Reddit contains valuable and unique public health information, adding to the growing evidence of the utility of social media for this type of public health surveillance, and stressing the importance of researchers continuing to have access to such data.^12–20,43^ With further development, social media surveillance systems could assist in identifying, monitoring, and predicting future drug epidemics.

## Materials and Methods

### Datasets

#### Reddit Opioid Comment Database

Reddit comments and their associated metadata from January 2006 to December 2022 were downloaded from PushShift.io and the Reddit API.^32^ Our initial drug lists came from Le *et al*.^37^ and the WHO’s Anatomical Therapeutic Chemical Classification System.^34^ Recognizing that Reddit users often use non-standard drug vocabularies, we used the RedMed word embedding model, which was trained on a health-oriented subset of Reddit, and which provides a lexicon of misspellings and synonyms for different drugs.^35, 36^ We initialized our opioid term list using terms linked to these two drug classes within the RedMed lexicon. To avoid the inclusion of ambiguous terms, we manually filtered the RedMed term list. We curated our opioid mention dataset from Pushshift data by selecting all comments that contained at least one of our opioid search terms.

#### Estimating Geo-Location and Creating a User Cohort

Neither Reddit nor the pushshift API provides location metadata for Reddit users, so we created a simple proxy to estimate user location. We extracted an initial set of location-specific subreddits from a Redditor curated list. We manually mapped these subreddits to their city, state, and region. We identified all Reddit users who posted at least once in a location-based subreddit during the observation period from 2010 to 2020 for all users. We then filtered out users who had posted in multiple location-based subreddits. Our final cohort consisted of all Reddit users who had posted in a single location-based subreddit at least once. We note that the inclusion criteria for this cohort was only our ability to get a proxy location for an individual user and was agnostic to whether they had ever mentioned opioids. We also selected a normalization strategy that minimized the impact inactive users had on the normalized rates, by normalizing our opioid discussion rates using the number of comments made by the cohort.

#### Opioid Receptor Activity-Based Classes

The opioids labeled as full agonists are morphine, codeine, oxycodone, pethidine, diamorphine, hydromorphone, levorphanol, methadone, fentanyl, sufentanyl, remifentanyl, tramadol, tapedolol, oxymorphone, and hydrocodone. The opioids labeled as partial agonists are buprenorphine, meptazinol, and loperamide. Mixed agonist opioids were nalorphine, pentazocine, nalbuphine, butorphanol, and dezocine. Opioid antagonists were naloxone, naltrexone, nalmefene, and diprenorphine. Heroin was maintained as an independent opioid class.

#### Opioid Synthesis-Based Classes

Opioids labeled as synthetic were tramadol, fentanyl, and meperidine. The class of natural and semi-synthetic opioids consisted of morphine, codeine, hydrocodone, oxycodone, oxymorphone, hydromorphone, naloxone, buprenorphine, and naltrexone. Heroin was maintained as an independent opioid class and the remaining opioids listed above and in Table 1 were grouped into the Opioid class. Methadone was excluded from these classes based on CDC classification.

### Benchmark datasets

#### CDC Vital Statistics Overdose Data

The CDC vital statistics unit has published trailing monthly drug overdose data which we collected for 2015-2022, for a subset of states and cities in the United States.^4^ The data is monthly 12-month trailing provisional monthly overdose deaths for several different opioid categories (all opioids, heroin, semi-synthetic opioids, and synthetic opioids). To convert the overdose data to overdose death rates, we normalized the opioid death counts by the size of the relevant population (i.e., location and year per 10,000 people) based on census data.

#### NFLIS Data

The NFLIS monitors drug use trends in different communities in the U.S., through reports from state and local forensic labs.^5^ These reports represent ∼98% of the data from the 1.5 million annual U.S. drug cases. The laboratory network includes data from 50 states and 104 local forensic labs. NFLIS provides drug identification rates for 25 drugs that are most commonly identified in national laboratory reports. This data is reported on a semi-annual basis and we collected data from 2014 to 2022 for the entire U.S. and for each region of the U.S.: south, midwest, west, and northeast. In order to convert the drug report data to drug report rates, we normalized the opioid report counts by the size of the relevant population (i.e. location and year per 100,000 people) based on census data.

### Calculation of Reddit and Benchmark Statistics Over Time

#### Summary Statistics and Unnormalized Opioid Mention Counts

We counted the total number of mentions in our opioid database for each drug to create summary statistics for the opioid discussion on Reddit. We counted the number of comments by month for each drug and opioid category and visualized these counts over time (Figure 1).

#### Normalized Rates: Comparing Reddit Comment Rates with CDC Overdose Data

To compare the Reddit opioid conversation with CDC overdose data, we calculated 12-month trailing opioid comment rates within our cohort for the geographies we covered, and for the categories the CDC reported: all opioids, heroin, semi-synthetic opioids, and synthetic opioids. The numerator for the Reddit opioid comment rate was the 12-month trailing total number of comments (based on the opioid category) for the geography (U.S., region, state, city) and the denominator was the 12-month trailing total number of comments made by users within our location cohort for the geographic area (U.S., region, state, city). We normalized these rates per 10,000 total comments.

We compared the 12-month trailing comment rates and the 12-month trailing overdose rates for each U.S. state. We grouped these rates and reported them for the entire U.S. and regions in the U.S. We visualized the 12-month trailing opioid comment rates and the 12-month trailing opioid overdose rates side by side for the entire U.S., and for different regions in the U.S. for 2015-2022 (Figure 3).

We compared the 12-month trailing comment rates and the 12-month trailing overdose rates for each category for the entire U.S., for regions in the U.S. using cross-correlation analysis, i.e. calculating correlations using different leading and lagging time intervals. We excluded the COVID-19 years, 2020 and 2021, from this analysis. We also performed an augmented Dickey-Fuller unit root test to evaluate the stationarity of the data. We did a subsequent cross-correlation analysis as described above after first or second-order differencing to achieve stationarity.

#### Normalized Rates: Comparing Reddit Comment Rates with NFLIS Drug Report Data

To compare the Reddit opioid conversation with the NFLIS drug report rates, we started by calculating the total number of semi-annual opioid comments for the geographies and select drugs: heroin, oxycodone, hydrocodone, buprenorphine, and fentanyl. The numerator for this normalized rate was the total number of semi-annual comments for the drug and geography (U.S., region), and the denominator for this was the total number of semi-annual comments made by users with location data for the geographic area (U.S., region). We normalized these rates per 100,000 total comments.

We then compared the semi-annual comment rate and the semi-annual drug report rate for these drugs for the entire U.S., and for NFLIS reporting regions in the U.S. We visualized the semi-annual drug comment rate and the semi-annual drug report rate side by side for the entire U.S., and for different regions in the U.S. for 2014 to 2022.

#### ARIMA Modeling

We fit our initial ARIMA model using 6-month lagged national CDC rates as described above from 2015 to 2022 using the python pmdarima auto_arima function.^38^ We then used the rolling forecast function with an initial training dataset spanning two years and a predictive time horizon of 1 month.^38^ We did not introduce seasonality in the model. We then calculated the average error of the predicted values against the true CDC monthly normalized overdose death rates (per 10,000 people) over time. We also fit an ARIMA model using the same CDC data but with real-time (1 month lagged) monthly Reddit comment rates (opioid comments per 10,000 total comments) incorporated as an exogenous variable. Finally, we fit the combined ARIMA model with the same parameters as with the Reddit-only model. To simulate real-world applications, we elected to lag the CDC data by 6 months, within the lag range reported by the CDC Vital Statistics Provisional Drug Overdose Death Counts.^42^ Performance was evaluated via Wilcox Signed Rank testing of absolute errors between predicted and observed values.

## Data Availability

The underlying data is not provided for ethical reasons.

## Acknowledgments

The authors would like to thank Shashanka Subrahmanya for his assistance. This work is supported by the NIH DA057598 and MH125702. Delaney Smith is supported by the Stanford Biochemistry Department and NSF GRFP 2019286895; Kieth Humphreys is supported by a Senior Research Career Scientist Award (RCS 04-141-3) from the Department of Veterans Affairs Health System Research Service; Doctors Eichstaedt, Altman, and Lembke are supported by the Stanford Institute for Human-Centered AI; Russ Altman is supported by the Chan Zuckerberg Biohub.

## Author Contributions

DAS analyzed the data, created some figures, and drafted the manuscript. AL developed the geo-location method used in this paper. AS processed raw Reddit data and created some figures and tables. TH analyzed the data. KH provided field expertise in addiction. MK provided expertise in statistical analysis and epidemiology. RBA assembled the research team and provided overall management of project. JCE edited the manuscript and provided guidance in analyzing, interpreting, and communicating the results.

## Code Availability

The underlying code for this study is not publicly available but may be made available to qualified researchers upon reasonable request from the corresponding author.

---

**Supplementary Figure 1:**
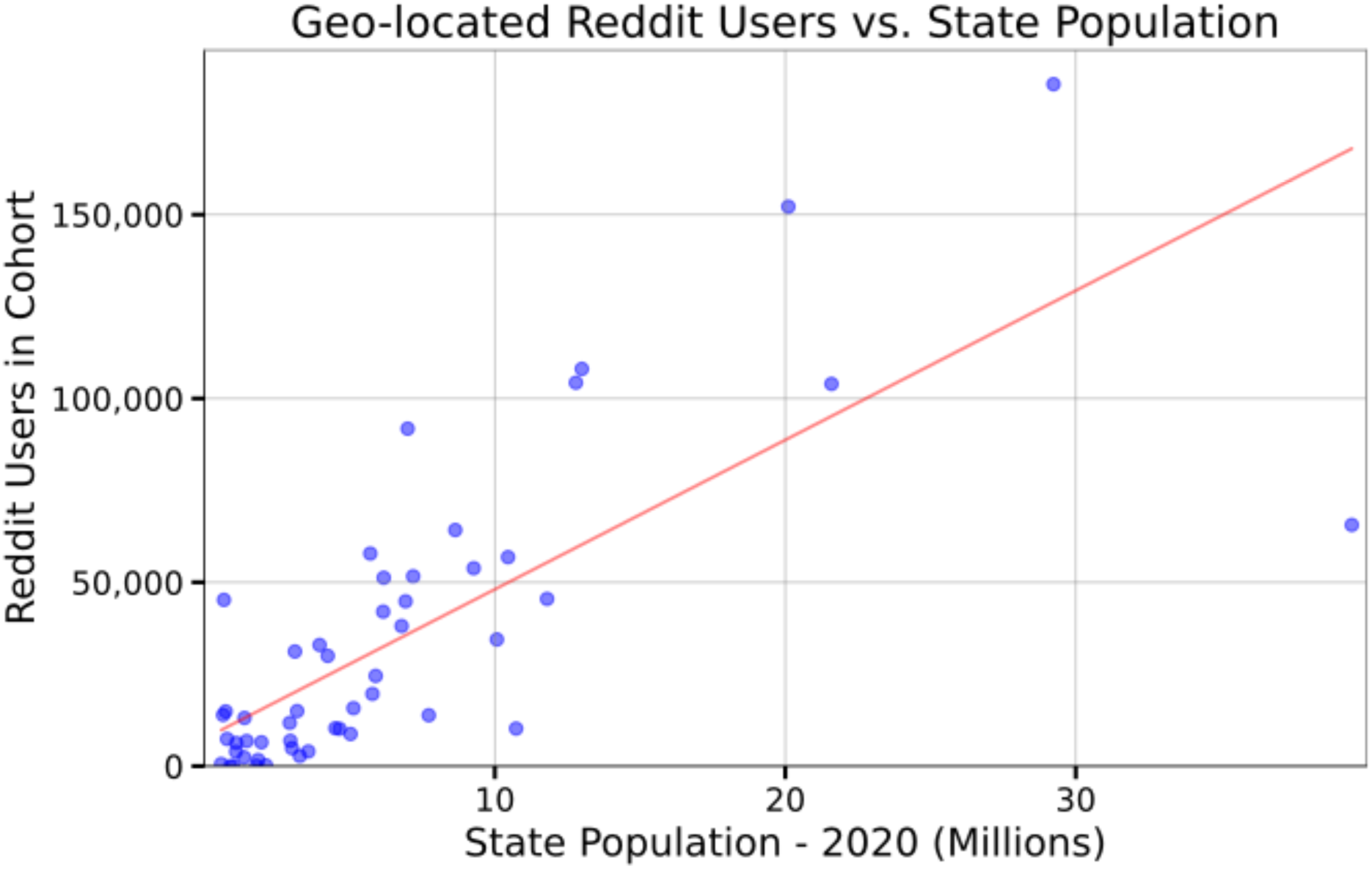
State populations in 2020 (x-axis) versus number of users assigned to that state in our cohort of geo-located users (Kendall’s Tau = 0.595). Four states had no observed users and were added to the plot with zero users.

**Supplementary Figure 2.**
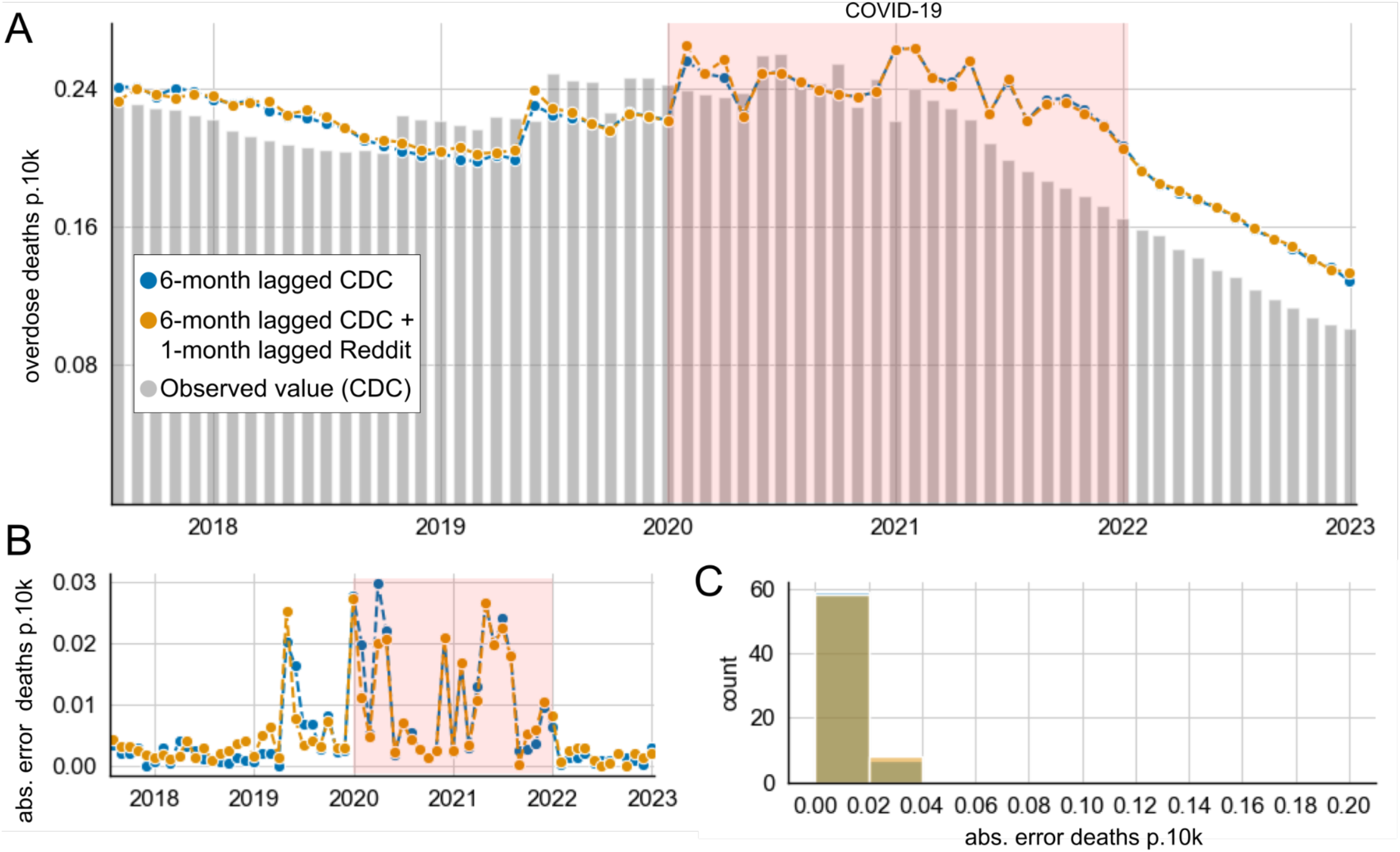
Autoregressive Integrated Moving Average (ARIMA) simulations for heroin. **A)** Predicted monthly overdose death rates per 10,000 people of rolling-origin forecast models are shown based on 1-month prediction horizons. Observed mortality is shown in grey, and monthly overdose death rates per 10,000 people as predicted by ARIMA models fitted on 6-month-lagged CDC overdose death (blue) are shown along with predictions from a model that additionally included 1-month-lagged Reddit data (orange). **B)** shows the absolute errors over time of the monthly overdose deaths per 10,000 people predicted by the lagged CDC model alone (in blue) and the combined CDC/Reddit model (in orange); **C)** shows the corresponding distributions. The combination Reddit/CDC model showed improved predictive accuracy for overdose death rates compared to the lagged CDC data alone, which did not rise to statistical significance (p = 0.200). The average absolute error for CDC alone was 0.0063, and 0.0062 for the combination model (in monthly overdose death rates per 10,000 people). In both models, we see notably decreased performance during the COVID years from 2020 until 2022. Before 2020, the combination model outperformed the CDC alone model (not significant at p = 0.105), with average absolute errors of 0.0048 and 0.0043 monthly normalized overdose death rates for CDC alone and CDC/Reddit, respectively.

**Supplementary Figure 3.**
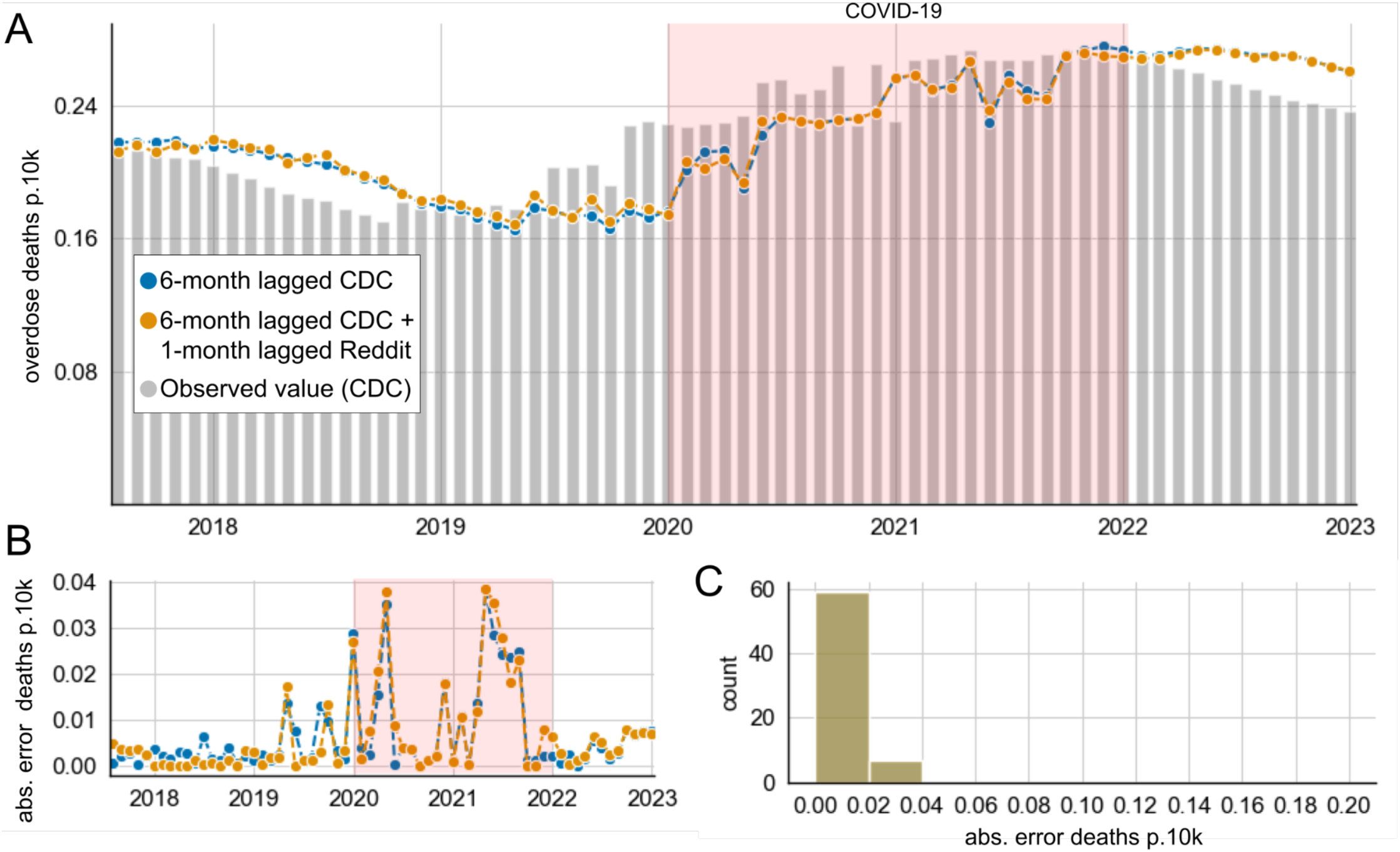
Autoregressive Integrated Moving Average (ARIMA) simulations for natural and semi-synthetic opioids. **A)** Predicted monthly overdose death rates per 10,000 people of rolling-origin forecast models are shown based on 1-month prediction horizons. Observed mortality is shown in grey, and monthly overdose death rates per 10,000 people as predicted by ARIMA models fitted on 6-month-lagged CDC overdose death (blue) are shown along with predictions from a model that additionally included 1-month-lagged Reddit data (orange). **B)** shows the absolute errors over time of the monthly overdose deaths per 10,000 people predicted by the lagged CDC model alone (in blue) and the combined CDC/Reddit model (in orange); **C)** shows the corresponding distributions. The distribution of absolute errors of the monthly overdose deaths per 10,000 people of the lagged CDC model alone (in blue) and the combined CDC/Reddit model (in orange). The Reddit/CDC model did not show improved accuracy over the CDC-only model in predicting overdose death rates (p = 0.865, Supplemental Figure 3). In both models, we see notably decreased performance during the COVID years from 2020 until 2022. Before 2020, the combination model outperformed the CDC alone model (not significant at p = 0.221), with average absolute errors of 0.00344 and 0.00341 for CDC alone and CDC/Reddit respectively.

